# Genomics of Post-Vaccination SARS-CoV-2 Infections During the Delta Dominated Second Wave of COVID-19 Pandemic, from Mumbai Metropolitan Region (MMR), India

**DOI:** 10.1101/2022.02.26.22271546

**Authors:** Kayzad Nilgiriwala, Pratibha Kadam, Grishma Patel, Ambreen Shaikh, Tejal Mestry, Smriti Vaswani, Shalini Sakthivel, Aruna Poojary, Bhavesh Gandhi, Seema Rohra, Zarir Udwadia, Vikas Oswal, Daksha Shah, Mangala Gomare, Kalpana Sriraman, Nerges Mistry

## Abstract

Vaccination against SARS-CoV-2 was launched in India in January 2021. Though vaccination reduced hospitalization and mortality due to COVID-19, vaccine breakthrough infections have become common. The present study was initiated in May 2021 to understand the proportion of predominant variants in post-vaccination infections during the Delta dominated second wave of COVID-19 in the Mumbai Metropolitan Region (MMR) in India and to understand any mutations selected in the post-vaccination infections or showing association with any patient demographics. We collected samples (n=166) from severe/moderate/mild COVID-19 patients who were either vaccinated (COVISHIELD/COVAXIN – partial/fully vaccinated) or unvaccinated, from a city hospital and from home isolation patients in MMR. A total of 150 viral genomes were sequenced by Oxford Nanopore sequencing (using MinION) and the data of 136 viral genomes were analyzed for clade/lineage and for identifying mutations in all the genomes. The sequences belonged to three clades (21A, 21I and 21J) and their lineage was identified as either Delta (B.1.617.2) or Delta+ (B.1.617.2 + K417N) or sub-lineages of Delta variant (AY.120/AY.38/AY.99). A total of 620 mutations were identified of which 10 mutations showed an increase in trend with time (May-Oct 2021). Associations of 6 mutations (2 in spike, 3 in orf1a and 1 in nucleocapsid) were shown with milder forms of the disease and one mutation (in orf1a) with partial vaccination status. The results indicate a trend towards reduction in disease severity as the wave progressed.

## INTRODUCTION

Severe Acute Respiratory Syndrome Coronavirus - 2 (SARS-CoV-2) – the causative agent of the novel coronavirus related disease (COVID-19), since its occurrence in late 2019, has been continuously evolving and leading to the emergence of new variants. In the light of the COVID-19 pandemic, the development of various types of vaccines was achieved at a remarkable speed, since vaccines are the most potent weapon for controlling the pandemic [1]. The national COVID-19 vaccination program in India was launched in January 2021, with approval for the vaccines – COVISHIELD (ChAdOx1-S/nCoV-19) (Serum Institute of India, Pune) and COVAXIN (BBV152) (Bharat Biotech, Hyderabad). Clinical trials have shown that both vaccines are reported to have >70% efficacy against symptomatic infections [2]. The immune response in post-vaccination infections seems to favour vaccine-escape mutations. Several studies in India have reported breakthrough infections and prevalence of variants of concern (VOC) in vaccinated and unvaccinated COVID-19 positive individuals [3]. The earliest evidence reported breakthrough infections in 19 out of 113 employees (16.8%) at a non-communicable disease healthcare facility in Delhi [4]. Another study reported the viral genomic characterization of six viral variants isolated from healthcare workers with breakthrough infections [5], whereas the study by Gupta et al. (2022) provided evidence that the majority of the breakthrough COVID-19 cases in India were infected with the Delta variant, with only 9.8% cases requiring hospitalization, and 0.4% fatalities, indicating that vaccination is helpful in reducing hospitalization and mortality [6].

Since the release of the first SARS-CoV-2 genome sequence in Jan 2020 [7], whole genome sequencing (WGS) has proved to be a powerful tool to identify genomic characteristics and for the development of newer diagnostics, genomic surveillance and contact tracing [8]. India experienced the second wave of COVID-19 during Apr-Jun 2021, with a peak in mid-May 2021 [9] whilst the first case of the Delta (B.1.617.2) was reported in Maharashtra in Mar 2021 [10], the first case of the Delta variant was detected in the Mumbai Metropolitan Region (MMR) only in May 2021. Moreover, during the peak of the second wave in May 2021 only 2,005 complete genomes of the Delta variant of SARS-CoV-2 were reported from India and only four genomes from MMR were reported in the Global Initiative on Sharing All Influenza Data (GISAID) database [11].

Hence, considering the limited genome sequences from the MMR and to obtain a better representation of viral genome sequences from the MMR towards understanding the prevalence of the predominant VOC in post-vaccination SARS-CoV-2infections, we conducted this study between May and Oct 2021. Clinico-demographic information from post-vaccination SARS-CoV-2 infections in MMR was collected in an attempt to understand the association (if any) of the variants with disease severity, vaccination status (Vaccinated / Unvaccinated), vaccine type (COVISHIELD/COVAXIN) and vaccination dose status (Partially vaccinated / Fully vaccinated). We performed phylogenetic and mutation analysis of the variants in our cohort to get insights into the genomic changes in the virus and to understand any longitudinal mutation-specific trends during the Delta dominant second wave.

## MATERIALS AND METHODS

### Patients and Sample Collection

This study was conducted between May and Oct 2021 by the Foundation for Medical Research (FMR), in collaboration with the Municipal Corporation of Greater Mumbai (MCGM), Mumbai and Breach Candy Hospital (BCH) Trust, Mumbai. Ethical clearance for the study was obtained from the Institutional Ethics Committees at FMR (FMR/IREC/C19/01/2021 and FMR/IREC/C19/02/2021) and BCH (P6/2021). A total of 166 patients were recruited as part of these collaborative studies, out of which 153 patients were post-vaccination COVID-19 cases and 13 patients were unvaccinated. All the COVID-19 cases were confirmed based on RT-PCR. Patients were recruited from the MMR, of whom 74 patients were hospitalized (fully vaccinated) and 92 patients were in home isolation (vaccinated with one dose, vaccinated with two doses or unvaccinated). For hospitalised patients, samples were collected by the tertiary care hospital doctors and in the case of home isolation participants, the samples were collected by a trained field researcher. Written informed consent was obtained from all the patients during recruitment regarding the collection of swab samples and patient metadata (Table 1). Naso/oro-pharyngeal swabs (n=74) and nasopharyngeal swabs (n=92) were collected from the patients, and samples were categorized based on disease severity defined as per ICMR guidelines dated 17^th^ May 2021 [12]. The vaccinated group had representation from both the adenoviral vaccine (COVISHIELD) and the inactivated whole virus vaccine (COVAXIN). The swabs were collected in a viral transport medium (VTM) and transported at 4° C to the FMR. A detailed case history was recorded at the time of recruitment.

**Table 1.** Patient demographics and clinical parameters.

| Characteristics | COVISHIELD |  | COVAXIN |  | Unvaccinated | Total cases |
| --- | --- | --- | --- | --- | --- | --- |
|  | Vaccinated with One Dose * | Vaccinated with Two Doses * | Vaccinated with One Dose * | Vaccinated with Two Doses * |  |  |
|  | n=21 | n=80 | n=2 | n=26 | n=7 | n=136 |
| **Age (years)** |  |  |  |  |  |  |
| Median | 35 | 64 | 24 | 41 | 34 | 54 |
| **Gender** |  |  |  |  |  |  |
| Male | 11 | 48 | 1 | 14 | 4 | 78(57%) |
| Female | 10 | 32 | 1 | 12 | 3 | 58(43%) |
| **Disease severity** |  |  |  |  |  |  |
| Mild | 19 | 62 | 2 | 23 | 7 | 113 |
| Moderate | 2 | 9 | 0 | 2 | 0 | 13 |
| Severe | 0 | 9 | 0 | 1 | 0 | 10 |
| **Symptoms** |  |  |  |  |  |  |
| Fever | 12 | 61 | 1 | 21 | 1 | 96(71%) |
| Cough | 18 | 45 | 3 | 20 | 6 | 92(68%) |
| Cold | 5 | 5 | 0 | 3 | 1 | 14(10%) |
| Chills | 1 | 7 | 0 | 9 | 1 | 18(13%) |
| Shortness of breath or difficulty in breathing | 1 | 17 | 0 | 3 | 0 | 21(15%) |
| Sore throat | 7 | 11 | 2 | 7 | 6 | 33(24%) |
| Loss of taste | 6 | 9 | 0 | 9 | 3 | 27(20%) |
| Loss of smell | 6 | 10 | 0 | 12 | 2 | 30(22%) |
| Headache | 3 | 13 | 0 | 2 | 1 | 19(14%) |
| Muscle aches | 6 | 15 | 1 | 6 | 0 | 28(21%) |
| Weakness | 0 | 10 | 0 | 1 | 0 | 11(8%) |
| Nausea | 0 | 2 | 0 | 1 | 0 | 3(2%) |
| Diarrhoea | 0 | 1 | 0 | 3 | 0 | 4(3%) |
| **Comorbidities** |  |  |  |  |  |  |
| Diabetes | 0 | 34 | 0 | 5 | 1 | 40(29%) |
| Hypertension | 0 | 36 | 0 | 1 | 0 | 37(27%) |
| Hypothyroid | 0 | 8 | 0 | 0 | 0 | 8(6%) |
| Cardiovascular disease | 0 | 13 | 0 | 2 | 0 | 15(11%) |

### RNA Isolation and RT-PCR

Viral RNA from the swab samples were isolated with QiaAmp viral RNA mini kit (Qiagen GmBH, Hilden, Germany) as per the manufacturer’s protocol. The RT-PCR was carried out in the Bio-Rad CFX96 (Bio-Rad Laboratories, California, USA), real-time PCR Detection System, and SARS-CoV-2 specific genes (N and ORF1) were detected using the COVIpath™ COVID-19 RT-PCR kit (Applied Biosystems-Invitrogen Bioservices India Pvt. Ltd.) as per the manufacturer’s protocol.

### cDNA Synthesis and Multiplex PCR

Subsequent to RT-PCR, RNA samples with Ct < 33 (150/166) were subjected to reverse transcriptase PCR to convert the SARS-CoV-2 RNA into cDNA for sequencing, using LunaScript RT SuperMix Kit (Cat no: E3010, NEB, Ipswich, USA) as per manufacturer’s protocol. The cDNA were amplified by ARTIC primers (version 3) for 27 initial samples and later by MIDNIGHT primers (Version: PCTR_9125_v110_revB_24Mar2021) for 123 samples. Two separate PCR amplification reactions (with pool-1 and pool-2 primers) for each sample were conducted in case of both protocols.

### Genome Sequencing by Oxford Nanopore

The amplified products (from pool-1 and pool-2) of respective samples were confirmed by agarose gel electrophoresis. Samples that showed amplification for both the primer pools were considered optimal for Nanopore library preparation and sequencing. Amplified DNA were subjected to barcoding and adapter ligation using either the rapid barcoding MIDNIGHT protocol or native barcoding ARTIC protocol [13, 14]. Quality control of sequencing was done by including one positive control and a no template control during each sequencing run. DNA libraries were sequenced using the SpotON flow cell (FLO-MIN106, Oxford Nanopore Technologies, UK) in a MinION MK1B sequencer using MinKNOW operating software for primary data acquisition (Oxford Nanopore Technologies, UK).

### Genomic Data Analysis

#### Raw Data Processing

Base-calling and demultiplexing were conducted using *Guppy (v5.0.17)* in high accuracy mode [15]. The resulting .fastq files were normalized by read length. The processed reads were filtered with the *field bioinformatics pipeline* (*v1.2.1*) [16]. Reads were aligned using *Minimap2 (v2.17)* [17] to the reference genome (MN908947.3). Variants were called using *Medaka (v.1.5.0)* [18] from the aligned reads and consensus FASTA were created using *samtools* (v1.14) [19]. *SnpEff (version latest core)* was used to annotate the discovered variants with reference strain NC_045512.

#### Lineage Analysis

The assembled SARS-CoV-2 genomes were assigned lineages using Phylogenetic Assignment of Named Global Outbreak LINeages (*PANGOLIN*) (*v3.1.17)* [20] with Ultrafast Sample Placement on Existing Trees (UShER) model *(v1.2.121)* [21].

#### Phylogenetic Analysis

The consensus FASTA files from the SARS-CoV-2 were aligned using *MAFFT (v7.489)* [22] and clustered using *Augur (v13.0.0)* [23]. Maximum likelihood trees were constructed with default parameters using *IQ-TREE* (*v2.1.3)* [24] and visualised with *Auspice (v2.32.1*) [25]. *Nextclade* [26] was used to assign clades to the sequences. A secondary tree was generated to compare the sequences from this study with whole genome sequences from the GISAID database from MMR between Dec 2020 and Oct 2021the GISAID database from MMR between Dec 2020 and Oct 2021.

#### Mutation Association Analysis

The association of each mutation (including lineage defining mutations) with clinical parameters such as disease severity (severe/moderate/mild), vaccination status (vaccinated/unvaccinated), vaccine type (COVISHIELD/COVAXIN) and vaccination dose (partially/fully vaccinated) were analyzed using the Chi-square test in *GraphPad Prism 6*.

To analyze the association of the mutations with clinical parameters, we considered four categories: disease severity (severe/moderate/mild), vaccination status (vaccinated/unvaccinated), vaccine type (COVISHIELD/COVAXIN) and vaccine dose (partial/fully vaccinated). Since the severe and moderate patients required oxygen supplementation, which indicated extensive lung pathology, we grouped the patient samples into two categories: severe/moderate and mild, to understand the association with disease severity (Table 2). In the dataset mentioned above, 620 mutations (including non-lineage and lineage defining mutations) were present from 136 patient samples. Each mutation was considered a feature in the analysis. Mutations found in less than 5% of patients that may not provide meaningful association were excluded from the analysis—any association of mutations with a clinical parameter having *p-value* < 0.05 was considered statistically significant.

**Table 2.** Distribution of samples based on clinical parameters (disease severity, vaccination status, vaccine type and vaccine dose status).

| Category | Sub-class 1 | Sub-class 2 | Total |
| --- | --- | --- | --- |
| Disease severity | Severe/Moderate<br>(n=23) | Mild<br>(n=113) | 136 |
| Vaccination status | Vaccinated<br>(129) | Unvaccinated<br>(7) | 136 |
| Vaccine Type | COVISHIELD<br>(n=101) | COVAXIN<br>(n=28) | 129 |
| Vaccine dose status | Partial vaccinated (23) | Fully vaccinated<br>(106) | 129 |

## RESULTS

### Demographics and Clinical Features of COVID-19 Patients

Of the 166 patients recruited, 16 patient samples were excluded due to high Ct values (Ct > 33). Of the 150 samples sequenced, the sequence data of 136 samples were considered for further analysis. Fourteen samples were excluded due to low sequence coverage either because the Ct values were high (n=13; Ct between 30-33) or there was a high number of ambiguous bases were observed (n=1). All further analyses were carried out on the genomic data of the 136 samples (Fig. 1). There were 57% (78/136) males and 43% (58/136) females among the analyzed patients. The patients’ ages ranged between 18 and 89 years, with a median age of 54 years. The demographics of 10 severe, 13 moderate and 113 mild cases are provided in Table 1.

**Fig. 1.**
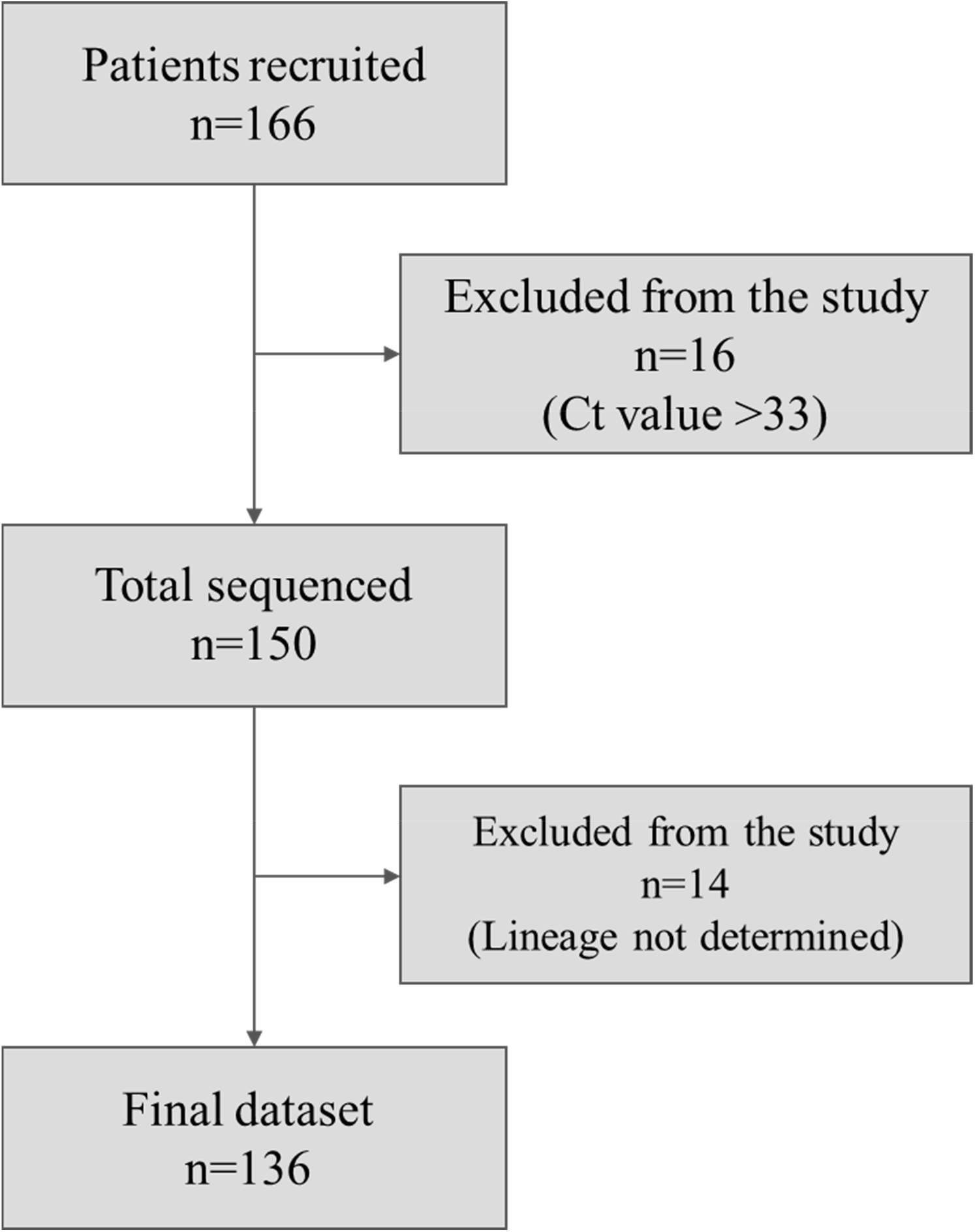
Schematic for sample selection in the study.

### Genome Sequencing Statistics

The sequence reads generated for the samples ranged between 0.1 and 1.17 million reads with a median sequence coverage (depth) of 5,327x. The GC content of the genomes ranged between 38 and 40%, and the sequenced genome length ranged from 28,918 to 29,890 bases. The genome coverage ranged from 96 to 100%. A range of 26-56 mutations per genome (SNP/Indels) were identified in the sequenced samples. The number of mutations ranged between 26 and 51 for mild cases, 31-56 for moderate between 31cases and 29-48 for severe between 29 and 48cases.

### Phylogeny of SARS-CoV-2 Genomes

The genomic data were analyzed for viral clades and lineages. The samples were found to be distributed in three clades based on the Nextclade classification, i.e., 21A (Delta), 21I (Delta), 21J (Delta). Six samples belonged to Clade 21A defined by positions 21618 (S: T19R), 23403 (S: D614G), 26767 (M: I82T), 28461 (N: D63G). Thirty-two samples belonged to Clade 21I defined by positions 5184 (nsp3:P1640L), 9891 (nsp4:3209V), 21618 (S: T19R), 22227 (A222V), 23403 (S:D614G), 26767 (M:I82T), 28461 (N:D63G). Ninety-eight samples belonged to Clade 21J defined by positions 11332 (nsp6:V3689V), 19220 (nsp14:A6319V), 21618 (S: T19R), 23403 (S:D614G), 26767 (M:I82T), 28461 (S: T19R).

Lineages were determined by PANGOLIN (Usher) interface, where 120 samples were found to be Delta (B.1.617.2), and 16 samples were Delta sub-lineages (AY-series and Delta+) (Table 3). A phylogenetic tree of the Clades 21A, 21I and 21J is shown in (Fig. 2). An additional phylogenetic tree was generated to compare genomes sequenced in this study with other sequenced genomes from MMR available in the GISAID database (Delta variants from MMR between May and Oct 2021) (Fig. 3). Sequences generated in this study and other MMR sequences (from GISAID) showed no grouping or presence of clusters in the phylogenetic profile. The Delta variants for samples collected during May-Aug 2021 belonged to either Clade 21A, 21I or 21J, whereas samples from Sep 2021 onwards belonged exclusively to 21J Clade. Additionally, the variants in Clade 21J showed an increase in the number of mutations compared to those in Clades 21A and 21I.

**Table 3.** Lineage distribution of Delta and its sub-lineages in the study.

| Lineage / sub-lineage | No. of samples |
| --- | --- |
| B.1.617.2 | 120 |
| AY.120 | 13 |
| AY.38 | 1 |
| AY.99 | 1 |
| B.1.617.2 + K417N (Delta+) | 1 |

**Fig. 2.**
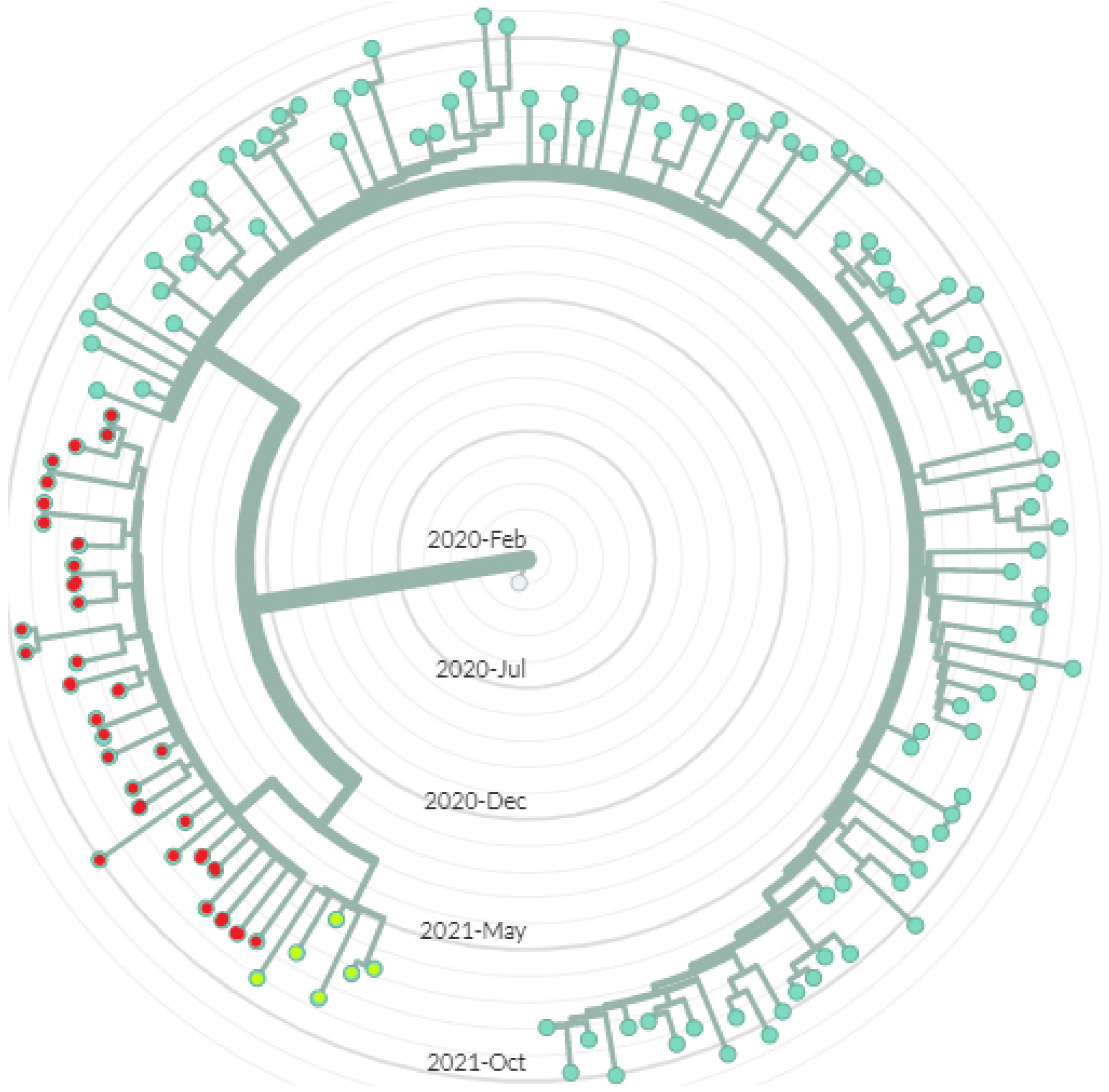
Phylogenetic tree representing 136 viral genome sequences from the study. The Clade 21A (parrot green), 21I (red) and 21J (green) are shown in a phylogenetic tree. The sequences from May to Aug 2021 belonged to either Clade 21A, 21I or 21J, and Sep 2021 onwards belonged to Clade 21J.

**Fig. 3.**
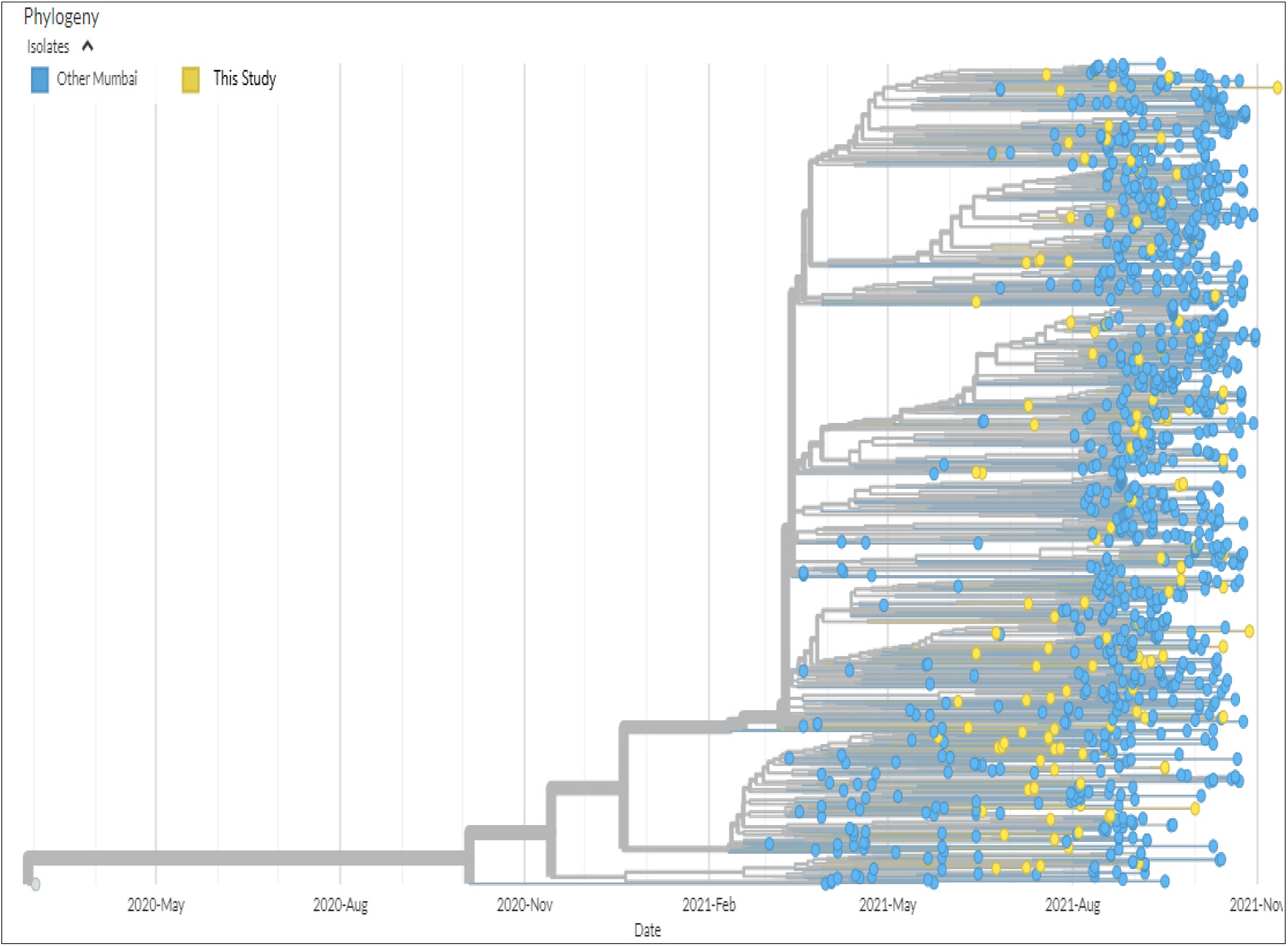
Phylogenetic tree of 136 viral genome sequences from this study (yellow) along with other viral genome sequences from MMR (blue) from the GISAID database (1,203 Delta variants and its sub-lineages).

### Association of Mutations with Clinical Parameters

A total of 620 mutations (SNP/Indels) were observed in 136 genomes in this study. We observed an increase in the frequency of 10 mutations over time (May to Oct 2021) (Fig. 4) which are as follows – 1) Orf1a: P2287S, 2) Orf1a: T3255I, 3) Orf1a: A1306S, 4) Orf1a: P2046L, 5) Orf1a: T3646A, 6) Orf1b: A6319V and 7) Orf7b: T40I, 8) S: G142D, 9) S: T95I, 10) N: G215C. These mutations showed a progressive increase in frequency (since Apr 2021), as also seen in the genome sequences available on the open-source database of COVID-19 resources and epidemiology data (https://outbreak.info/) (Fig. 4). In our study, these mutations were prevalent in over 70% of Delta variants, including Delta sub-lineages.

**Fig. 4.**
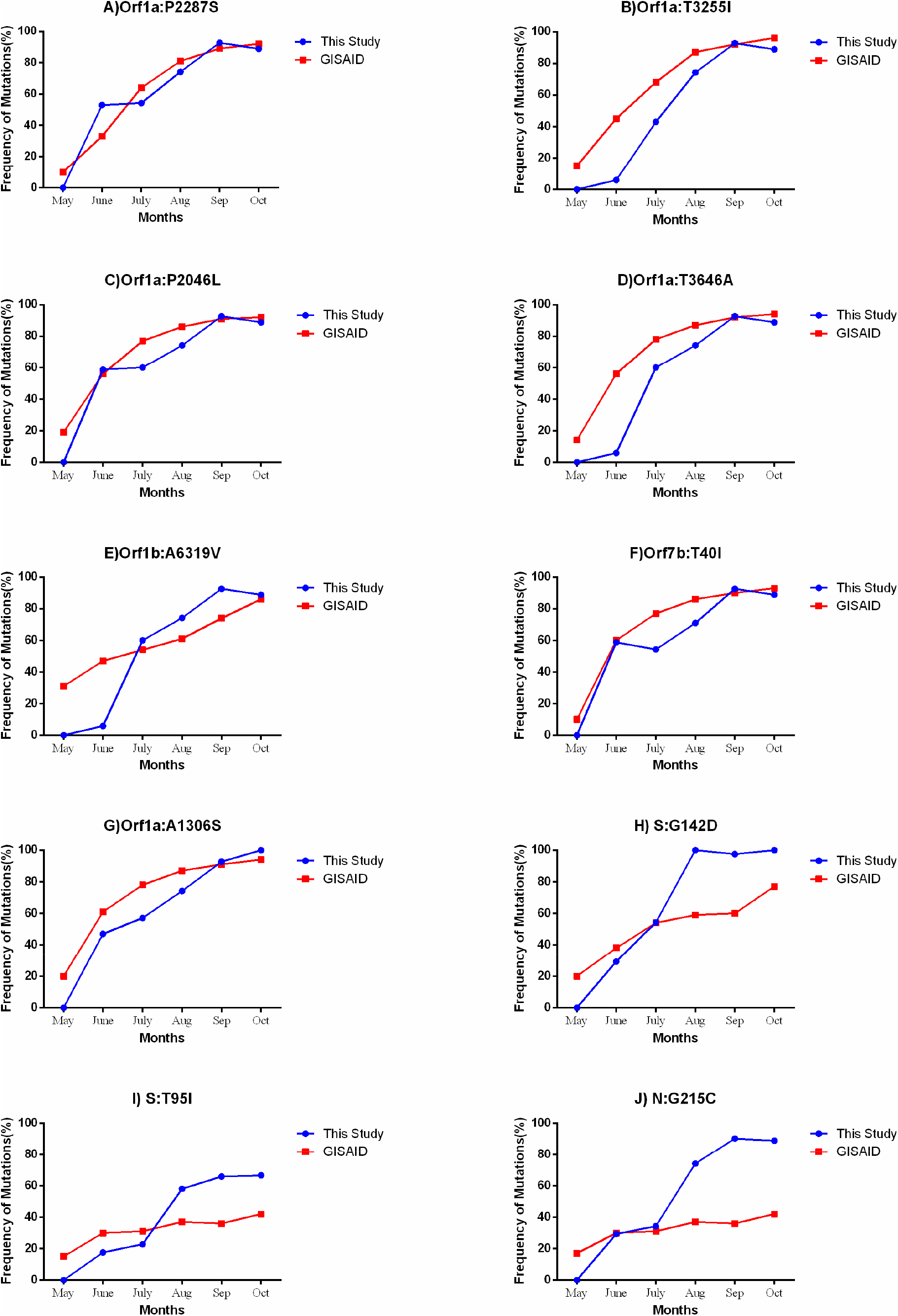
Mutations showing an increase in frequency from May to Oct 2021. A) Orf1a: P2287S B) Orf1a: T3255I C) Orf1a: P2046L, D) Orf1a: T3646A, E) Orf1b: A6319V F) Orf7b: T40I G) Orf1a: A1306S H) S: G142D I) S: T95I J) N: G215C. The graphs were generated using GraphPad Prism 6. The trendline in blue indicates the proportion of the respective mutations from this study, whereas the red trendline indicates the proportion of the respective mutations in the GISAID repository.

To analyze the association of the mutations with clinical parameters, we considered four categories: disease severity (severe/moderate/mild), vaccination status (vaccinated/unvaccinated), vaccine type (COVISHIELD/COVAXIN) and vaccine dose (partial/fully vaccinated). In the dataset, as mentioned above, 620 mutations were present from 136 patient samples, and each mutation was considered a feature in the analysis.

A comparison between the presence of mutations with the disease severity categories (severe/moderate vs mild) showed an association of six mutations (2 in S gene, 3 in orf1a and 1 in N gene) (Table 4) with mild cases (*p-value* < 0.05). Three mutations (1 in spike – G142D and 2 in orf1a - P2287S and T3255I) out of the six mutations showing association with the mild cases were also observed to have an increased frequency over time (between May-Oct 2021) (Fig. 4), and with two mutations found to be lineage defining for Delta (Table 4).

**Table 4.**
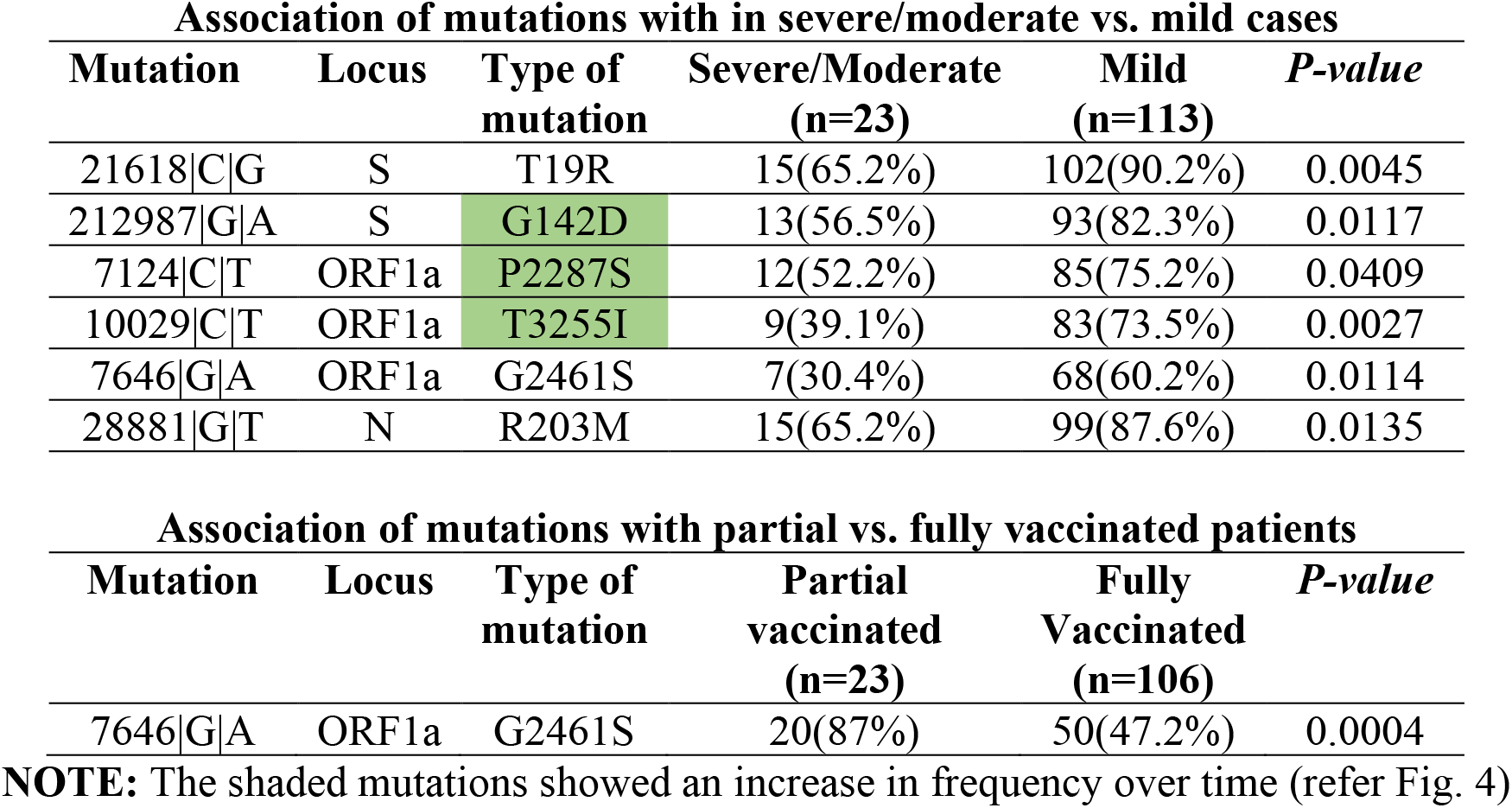
Mutations showing association with disease severity and vaccine dose.

| **Association of mutations with in severe/moderate vs. mild cases** |  |  |  |  |  |
| --- | --- | --- | --- | --- | --- |
| **Mutation** | **Locus** | **Type of mutation** | **Severe/Moderate (n=23)** | **Mild (n=113)** | **P-value** |
| 21618\|C\|G | S | T19R | 15(65.2%) | 102(90.2%) | 0.0045 |
| 212987\|G\|A | S | G142D | 13(56.5%) | 93(82.3%) | 0.0117 |
| 7124\|C\|T | ORF1a | P2287S | 12(52.2%) | 85(75.2%) | 0.0409 |
| 10029\|C\|T | ORF1a | T3255I | 9(39.1%) | 83(73.5%) | 0.0027 |
| 7646\|G\|A | ORF1a | G2461S | 7(30.4%) | 68(60.2%) | 0.0114 |
| 28881\|G\|T | N | R203M | 15(65.2%) | 99(87.6%) | 0.0135 |

| **Association of mutations with partial vs. fully vaccinated patients** |  |  |  |  |  |
| --- | --- | --- | --- | --- | --- |
| **Mutation** | **Locus** | **Type of mutation** | **Partial vaccinated (n=23)** | **Fully Vaccinated (n=106)** | **P-value** |
| 7646\|G\|A | ORF1a | G2461S | 20(87%) | 50(47.2%) | 0.0004 |

**NOTE:** The shaded mutations showed an increase in frequency over time (refer Fig. 4)

A comparison between the presence of mutations in partially vaccinated and fully vaccinated patients showed an association with a single mutation in orf1a with partially vaccinated patients (*p-value* < 0.001) (Table 4). A comparison between vaccinated and unvaccinated patients and between vaccine types (COVISHIELD vs COVAXIN) did not show any statistically significant association.

## DISCUSSION

Overall it is evident that COVID-19 is more lethal in unvaccinated people than those fully vaccinated, with vaccines providing the necessary protection from the causative virus [27]. Although vaccines can help reduce the severity of the disease in patients, they do not stop infection and do not reduce transmission. With the ever-changing virus during a pandemic, it becomes necessary to understand the pattern of its transformation, about vaccination rates and disease severity. The world has seen a repertoire of VOCs during this pandemic leading to the realization of the important role of different variants in manifesting varying levels of disease severity and transmission, creating abrupt surges (or waves) in the number of cases in turn increasing transmissions in various countries. In order to get insights into the newly acquired mutations in Delta variants from MMR, we screened for the mutations that showed an increase in trend from May to Oct 2021. We found that 10 mutations showed increased frequency with time. The comparison of mutations with the Mumbai sequencing data (available on the GISAID) indicated that these mutations appear to stabilize over time (Fig. 4) and may have potential implications in tuning transmission and infection levels of the virus, possibly leading to the generation of milder viral variants in the future.

We found the association of six mutations with mild cases (Table 4); three of those mutations were also observed to have an increased frequency with time (between May and Oct 2021) (Fig. 4). We found two mutations in the spike protein (T19R and G142D) (Fig. 5) associated with mild disease. The spike mutation T19R is a lineage-defining mutation found in most Delta variants. In contrast, the G142D mutation has been observed in 49% Delta variants and 69% Delta Plus variants in the global database [28]. We found 117/136 (86%) samples with co-occurrence of the T19R and G142D mutations in the spike; these mutations are known to change the supersite epitope that binds the N-terminal domain directed antibodies leading to immune evasion [28, 29]. Moreover, 41% of our samples had co-occurrence of T95I and G142D mutations in the spike. The co-occurrence of these mutations is reported to increase the viral load significantly [30]. The increase in viral load due to increased transmission fitness or immune escape may possibly lead to a tradeoff with disease severity leading to a milder disease. The presence of orf1a: T3255I, S: T95I and S: G142D mutations in all the Omicron (B.1.1.529) variants – a variant supposed to lead to a milder disease reiterates the association of these mutations with reduced severity [31, 32].

**Fig. 5.**
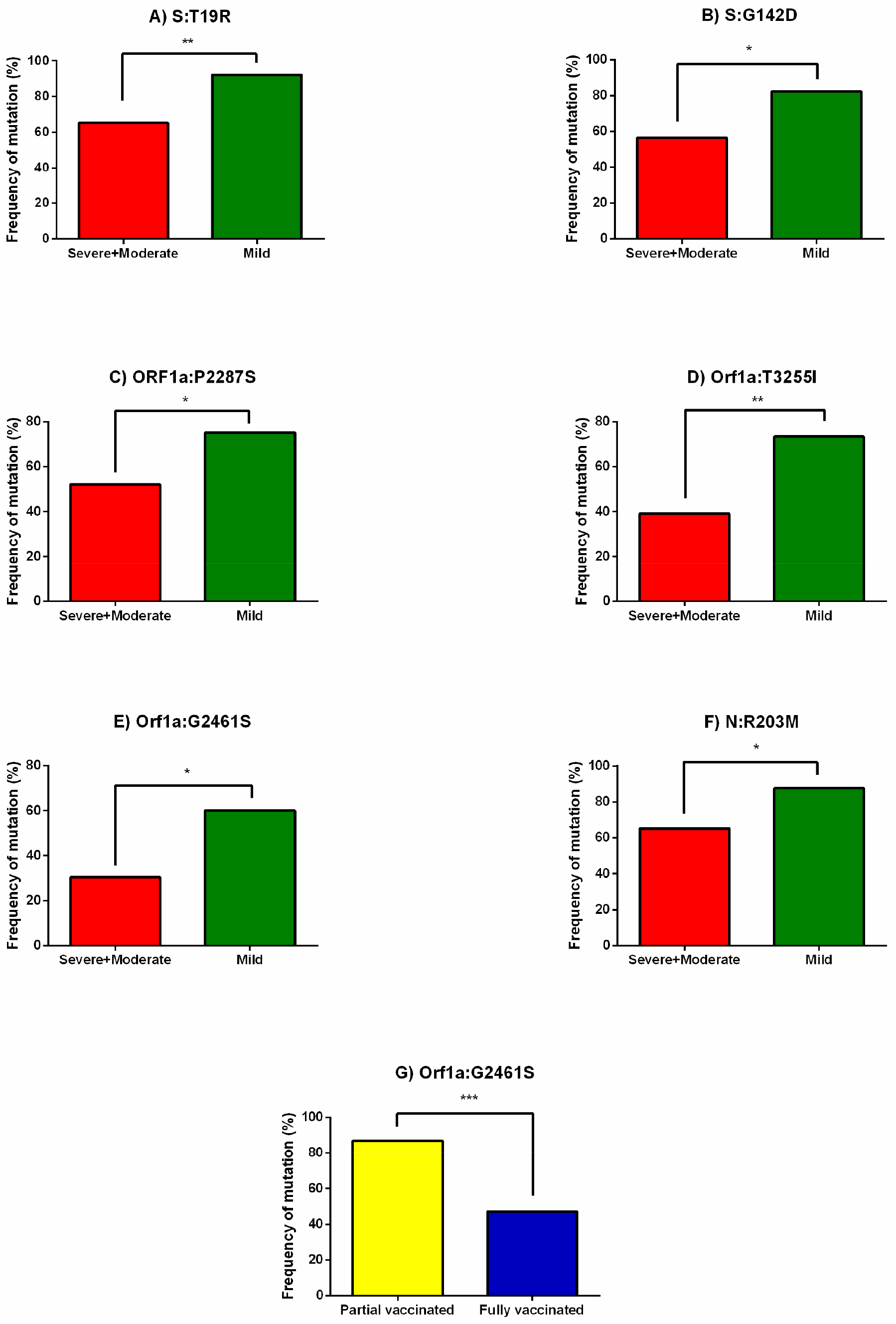
Association of mutations with disease severity and vaccination status (partially/fully vaccinated). Six mutations showed association with mild cases: A) S: T19R B) G142D C) Orf1a: P2287S D) Orf1a: T3255I E) orf1a: G2461S F) N: R203M. One mutation G) orf1a: G2461S showed association with partially vaccinated cases.

We also found an association of three mutations in orf1a (P2287S, T3255I and G2461S) with the mild disease, of which two mutations in orf1a (P2287S and T3255I) were also observed to increase with time (Figs. 4 and 5). The orf1a: P2287S mutation is present in the nsp3 Plpro papain-like protease whose function is to block host innate immune response and promote cytokine expression [33]. The increasing frequency of orf1a: P2287S mutation and its association in mild cases could possibly indicate an enhanced host immune response and reduced cytokine expression (potentially mitigating cytokine storm), leading to less severe disease. Another mutation in orf1a associated with mild disease in nsp3 PLpro papain-like protease (orf1a: G2461S) could possibly play a similar role in causing less severe disease by enhancing host immune response and mitigation of cytokine storm. Interestingly, the orf1a: G2461S mutation also showed association in partially vaccinated cases indicating selection of the mutation after the first dose of vaccine. However, the limitation of the meagre number of partially vaccinated patients (n=23) in this study should be noted before drawing any conclusion. The third mutation in orf1a (T3255I) is present in the nsp4 protein, whose function is to produce double-membrane vesicles required to form a replication-transcription complex [33]. This mutation may play a role in reducing the formation of active replication-transcription complexes in the host cell, in turn reducing the viral load and explaining its association with the milder disease form.

In contrast, another mutation in the nucleocapsid protein (N: R203M) was observed to associate with the milder form of the disease (Fig. 5). It has already been reported that the N: R203M mutation leads to increased packaging of the viral RNA genome producing a 50-fold higher viral load [34]. The association of the N: R203M mutation with milder disease could possibly explain the increase in viral load due to an increase in variant transmission fitness, leading to a tradeoff with disease severity. The increase in the frequency of the mutations orf1a (T3255I) (leading to a potential decrease in viral load) and N: R203M (leading to a possible increase in viral load) in the recent Delta variants and their association with milder disease indicates their significant role in determining the transmission fitness of the newer variants and tradeoff with disease severity.

A continued global effort towards ever expanding viral genome sequencing will help to further the knowledge on potential novel mutations of interest and to understand the evolution of the virus regarding its behaviour in transmission and epidemiology.

## Data Availability

The genomic data generated in this study can be freely downloaded from the GISAID repository (GISAID accession numbers EPI_ISL_8164107 - EPI_ISL_8164235 and EPI_ISL_8173172 - EPI_ISL_8173178).

https://www.gisaid.org

## DATA AVAILABILITY

The genomic data generated in this study can be freely downloaded from the GISAID repository [11] (GISAID accession numbers EPI_ISL_8164107 - EPI_ISL_8164235 and EPI_ISL_8173172 - EPI_ISL_8173178).

## ACKNOWLEDGEMENTS

The authors thank all the participants of this study and thank Mr. Nadir Godrej, Mr. Aditya Berlia, Mr. Rakesh Agarwal, Mr. Pranav Kothari, Mr. Sandeep Chopra, Mr. Anantnarayan Sunderesan and others for their donations for conducting the project. The authors also thank Dr. Chandrakant Pawar, Medical Superintendent, Kasturba Hospital for Infectious Diseases for his support towards the study. The contribution of Ms. Niharika Shinde, the field researcher is appreciated for enrolling the patients and sample collection.

## AUTHOR CONTRIBUTIONS

**Conceptualization** – KN, AP, ZU, VO, KS, NM

**Data Curation** – KN, PK, GP, TM, AS, SV, AP, BG, SR, KS

**Formal Analysis** – KN, PK, AS, SV, KS

**Funding Acquisition** – KS, NM

**Investigation** – PK, GP, TM, AS, SV, SS, BG, SR

**Methodology** – KN, AP, KS, NM

**Project Administration** – KN, PK, GP, TM, AS, AP, KS, NM

**Resources** - KN, PK, AP, ZU, VO, DS, MG, NM

**Software** – KN, PK

**Supervision** – KN, AS, AP, ZU, DS, KS, NM

**Validation** – KN, PK, GP, TM, AS, SV

**Visualization** – KN, PK, GP, TM

**Writing – Original Draft Preparation** – KN, PK, GP, TM

**Writing – Review and Editing** – KN, PK, GP, TM, AS, SV, AP, ZU, DS, KS, NM

## REFERENCES

1. Pollard, A.J. and E.M. Bijker, A guide to vaccinology: from basic principles to new developments. Nature reviews. Immunology, 2021. 21(2): p. 83–100.

2. Sharma, K., et al., Vaccines for COVID-19: Where do we stand in 2021? Paediatric respiratory reviews, 2021. 39: p. 22–31.

3. Thangaraj, J.W.V., et al., Predominance of delta variant among the COVID-19 vaccinated and unvaccinated individuals, India, May 2021. The Journal of infection, 2022. 84(1): p. 94–118.

4. Tyagi, K., et al., Breakthrough COVID19 infections after vaccinations in healthcare and other workers in a chronic care medical facility in New Delhi, India. Diabetes & metabolic syndrome, 2021. 15(3): p. 1007–1008.

5. Philomina J, B., et al., Genomic survey of SARS-CoV-2 vaccine breakthrough infections in healthcare workers from Kerala, India. The Journal of infection, 2021. 83(2): p. 237–279.

6. Gupta, N., et al., Clinical characterization and Genomic analysis of COVID-19 breakthrough infections during second wave in different states of India. 2021, Cold Spring Harbor Laboratory.

7. Wu, F., et al., A new coronavirus associated with human respiratory disease in China. Nature, 2020. 579(7798): p. 265–269.

8. Grifoni, A., et al., The proximal origin of SARS-CoV-2. Nature medicine, 2020. 26(4): p. 450–452.

9. Yang, W. and J. Shaman, COVID-19 pandemic dynamics in India, the SARS-CoV-2 Delta variant, and implications for vaccination. medRxiv, 2021.

10. Cherian, S., et al., Convergent evolution of SARS-CoV-2 spike mutations, L452R, E484Q and P681R, in the second wave of COVID-19 in Maharashtra, India. 2021, Cold Spring Harbor Laboratory.

11. GISAID Database. Available from: https://www.gisaid.org.

12. Clinical Guidance for Management of Adult COVID-19 patients (ICMR). 2021.

13. Quick, J. nCoV-2019 sequencing protocol v3 (LoCost) V.3. 2020; Available from: https://www.protocols.io/view/ncov-2019-sequencing-protocol-v3-locost-bh42j8ye.

14. Nilgiriwala, K., et al., Genome Sequences of Five SARS-CoV-2 Variants from Mumbai, India, Obtained by Nanopore Sequencing. Microbiology Resource Announcements, 2021. 10(15): p. e00231–21.

15. Wick, R.R., L.M. Judd, and K.E. Holt, Performance of neural network basecalling tools for Oxford Nanopore sequencing. Genome biology, 2019. 20(1): p. 129–129.

16. The ARTIC field bioinformatics pipeline. Available from: https://github.com/artic-network/fieldbioinformatics.

17. Li, H., Minimap2: pairwise alignment for nucleotide sequences. Bioinformatics (Oxford, England), 2018. 34(18): p. 3094–3100.

18. Nanopore/medaka. Available from: https://github.com/nanoporetech/medaka.

19. Li, H., et al., The Sequence Alignment/Map format and SAMtools. Bioinformatics (Oxford, England), 2009. 25(16): p. 2078–2079.

20. O’Toole, Á., et al., Assignment of epidemiological lineages in an emerging pandemic using the pangolin tool. Virus evolution, 2021. 7(2): p. veab064–veab064.

21. Turakhia, Y., et al., Ultrafast Sample placement on Existing tRees (UShER) enables real-time phylogenetics for the SARS-CoV-2 pandemic. Nature Genetics, 2021. 53(6): p. 809–816.

22. Katoh, K. and D.M. Standley, MAFFT multiple sequence alignment software version 7: improvements in performance and usability. Molecular biology and evolution, 2013. 30(4): p. 772–780.

23. Huddleston, J., et al., Augur: a bioinformatics toolkit for phylogenetic analyses of human pathogens. Journal of open source software, 2021. 6(57): p. 2906.

24. Chernomor, O., A. von Haeseler, and B.Q. Minh, Terrace Aware Data Structure for Phylogenomic Inference from Supermatrices. Systematic biology, 2016. 65(6): p. 997–1008.

25. McBroome, J., et al., A Daily-Updated Database and Tools for Comprehensive SARS-CoV-2 Mutation-Annotated Trees. Molecular biology and evolution, 2021. 38(12): p. 5819–5824.

26. Aksamentov, I., et al., Nextclade: clade assignment, mutation calling and quality control for viral genomes. Journal of Open Source Software, 2021. 6(67): p. 3773.

27. Dyer, O., Covid-19: Unvaccinated face 11 times risk of death from delta variant, CDC data show. BMJ, 2021: p. n2282.

28. Kannan, S.R., et al., Evolutionary analysis of the Delta and Delta Plus variants of the SARS-CoV-2 viruses. Journal of autoimmunity, 2021. 124: p. 102715–102715.

29. Planas, D., et al., Reduced sensitivity of SARS-CoV-2 variant Delta to antibody neutralization. Nature, 2021. 596(7871): p. 276–280.

30. Shen, L., et al., Spike Protein NTD mutation G142D in SARS-CoV-2 Delta VOC lineages is associated with frequent back mutations, increased viral loads, and immune evasion. 2021, Cold Spring Harbor Laboratory.

31. Micheli, V., et al., First identification of the new SARS-CoV-2 Omicron variant (B.1.1.529) in Italy. Clin Infect Dis, 2022.

32. Madhi, S.A., et al., Population Immunity and Covid-19 Severity with Omicron Variant in South Africa. N Engl J Med, 2022.

33. Perez-Gomez, R., The Development of SARS-CoV-2 Variants: The Gene Makes the Disease. Journal of developmental biology, 2021. 9(4): p. 58.

34. Syed, A.M., et al., Rapid assessment of SARS-CoV-2–evolved variants using virus-like particles. Science, 2021. 374(6575): p. 1626–1632.

